# Machine learning across multiple imaging and biomarker modalities in the UK Biobank improves genetic discovery for liver fat accumulation

**DOI:** 10.1101/2024.01.06.24300923

**Authors:** Hari Somineni, Sumit Mukherjee, David Amar, Jingwen Pei, Karl Guo, David Light, Kaitlin Flynn, insitro Research Team, Chris Probert, Thomas Soare, Santhosh Satapati, Daphne Koller, David J. Lloyd, Colm O’Dushlaine

## Abstract

Metabolic dysfunction-associated steatotic liver disease (MASLD), liver with more than 5.5% fat content, is a leading risk factor for chronic liver disease with an estimated worldwide prevalence of 30%. Though MASLD is widely recognized to be polygenic, genetic discovery has been lacking primarily due to the need for accurate and scalable phenotyping, which proves to be costly, time-intensive and variable in quality. Here, we used machine learning (ML) to predict liver fat content using three different data modalities available in the UK Biobank: dual-energy X-ray absorptiometry (DXA; n = 46,461 participants), plasma metabolites (n = 82,138), and anthropometric and blood-based biochemical measures (biomarkers; n = 262,927). Based on our estimates, up to 29% of participants in UKB met the criteria for MASLD. Genome-wide association studies (GWASs) of these estimates identified 15, 55, and 314 loci associated with liver fat predicted from DXA, metabolites and biomarkers, respectively, totalling 321 unique independent loci. In addition to replicating 9 of the 14 known loci at genome-wide significance, our GWASs identified 312 novel loci, significantly expanding our understanding of the genetic contributions to liver fat accumulation. Genetic correlation analysis indicated a strong correlation between ML-derived liver fat across modalities (*r*_g_ ranging from 0.85 to 0.96) and with clinically diagnosed MASLD (*r*_g_ ranging from 0.74 to 0.88), suggesting that a majority of the newly identified loci are likely to be relevant for clinical MASLD. DXA exhibited the highest precision, while biomarkers demonstrated the highest recall, respectively. Overall, these findings demonstrate the value of leveraging ML-based trait predictions across orthogonal data sources to improve our understanding of the genetic architecture of complex diseases.

## Introduction

Metabolic dysfunction-associated steatotic liver disease (MASLD), characterized by liver fat content exceeding 5.5% (hepatic steatosis), is a leading risk factor for end-stage liver disease, affecting more than 30% of the global population^1^. While MASLD is moderately heritable (∼39%)^2^, and is widely recognized to be polygenic, to date genome-wide association studies (GWASs) have identified only about 15 replicable loci harboring common genetic variants^3,4^, which stands in stark contrast to other common diseases with similar prevalence and heritability^5,6^. Genetic discovery in MASLD has been lacking in large part because of limited sample sizes (the number of cases in particular), underdiagnosis, and the methodologic challenge of accurately determining the phenotype in terms of the presence or absence of disease. For instance, the largest GWAS meta-analysis of clinical MASLD involved 9,491 cases, whereas the GWAS for a disease with a comparable prevalence and heritability, such as type 2 diabetes, included 180,834 cases^6^. Similarly, previous studies have demonstrated that, while fewer than 2% of the participants in the UK Biobank (UKB) have received the diagnosis (ICD9/10) for MASLD, 17% of the participants met the imaging-based criteria for MASLD, defined as liver fat content greater than 5.5%^7^. Finally, quantitative trait GWASs offer enhanced statistical power over binary-label GWASs, since the use of continuous data captures a broader range of phenotypic variation, minimizes information loss inherent in binary categorization, enables the study of intermediate phenotypes, and enhances accuracy by avoiding misclassification issues. These challenges underscore the need for larger and better phenotyped datasets to better understand the genetic architecture of liver fat accumulation, and by extension, hepatic steatosis related pathologies.

Magnetic resonance imaging (MRI)-derived proton density fat fraction (PDFF) is a widely used non-invasive method for accurately quantifying liver fat, allowing the detection of the presence and degree of hepatic steatosis^8^. While highly effective, the computation of PDFF is costly and time-intensive^9^. Recent studies by Langner *et al.*^10^ and Haas *et al.*^7^ have demonstrated the utility of machine learning (ML) techniques in predicting liver fat content from MRI, achieving remarkably high predictive performance with R^2^ values exceeding 0.90. However, the high costs, limited availability, and logistical challenges associated with MRI scanning present significant barriers for the large-scale measurements of liver fat needed for well-powered GWASs^11^. Therefore, there is a need for alternative data modalities and methods that are easily scalable and cost-effective for the quantification of liver fat content.

Dual-energy X-ray absorptiometry (DXA) has long been used as a valuable imaging modality for estimating bone density and body composition, encompassing measurements of fat and muscle mass^12^. Although DXA scans may not offer the same level of detailed information as MRI, they present distinct advantages: they are generally more cost-effective, widely available, and easier to obtain^13^. While there are reports suggesting the limited utility of DXA in quantifying liver fat content, these studies are constrained by factors such as small sample size, training without test sets, and the absence of reliable ground-truth measures, such as PDFF^14,15,16^.

In an alternative approach, anthropometric and blood-based biochemical measures (collectively referred to as biomarkers throughout this study), have recently been used to gain insights into the genetic underpinnings of MASLD. For instance, Miao *et al*.^17^ used ML on a set of biomarkers to construct a synthetic MASLD case-control cohort in the UKB. This cohort of 28,396 imputed MASLD cases and 108,652 controls was then used in a GWAS, reporting 94 loci. However, it is important to note that their approach comes with certain limitations, including a constrained sample size, and the use of binary labels. Furthermore, the reported associations were not thoroughly characterized in terms of their relevance to clinically diagnosed MASLD. Similar to biomarkers, metabolomics - for example, lipidomics-based algorithms - have been developed for the diagnosis of MASLD, but their performance has been demonstrated only on small cohorts^18^. Some of these limitations could be addressed by increasing sample size and/or by measuring a wider metabolic profile in each patient^19^.

In this study, we leverage ML on three different data modalities available in the UKB - DXA, metabolites and biomarkers - to predict liver fat content, with varying phenotypic accuracies and sample sizes. Our ML-derived predictions of liver fat content quantify liver fat as a continuous trait and/or indicate binary classification of MASLD. We conduct GWAS of ML-derived liver fat content, separately, within each of our data modalities, alongside PDFF, significantly expanding the genetic landscape of liver fat accumulation. Using pairwise genetic correlations, we demonstrate that a majority of GWAS associations are likely relevant to clinically diagnosed MASLD. Furthermore, we perform a comparative analysis of genetic loci across modalities, demonstrating the robustness of our findings. Our results underscore the value of ML approaches, highlighting their ability to leverage diverse data modalities as part of a comprehensive strategy for novel target discovery.

## Methods

### Data used

#### The UKB study

This research was conducted using the UKB resource under the approved application number 51766. A full description of the UKB study design is presented elsewhere^20^. Briefly, the UKB is a population-based, prospective cohort consisting of approximately half a million individuals between the ages of 40-69 with paired genetic and phenotypic information^21^. These participants were enrolled between 2006 and 2010 from multiple sites across the United Kingdom.

#### The UKB imaging initiative

The UKB initiated the largest multi-modal imaging study in 2014, with the goal of collecting imaging data from 100,000 previously enrolled participants. As part of this initiative, a significant proportion of the targeted 100,000 participants underwent brain, cardiac and abdominal MRI, DXA, and carotid ultrasound between 2014 and 2019^22^.

#### Metabolic markers

A panel of 249 metabolites (168 measures in absolute levels and 81 ratio measures) including lipoprotein lipids, fatty acids, and small molecules such as amino acids, ketones, and glycolysis metabolites, were measured using the Nightingale Health nuclear magnetic resonance (NMR) platform from blood samples collected at baseline from 118,461 participants (first data release, made available to approved UKB researches in 2021)^24^. In addition to baseline measurements, repeat assessments from a subset of about 1,500 participants were also included in the same data release. Details of the Nightingale Health NMR platform have been described previously^25,26^. The quality control protocol used to process this dataset has been described in detail^24^.

#### Biochemistry markers

A total of 34 biochemical markers were measured in biological samples (blood, urine and saliva) collected at baseline from all 500,000 participants^27,28^. These markers were selected based on their relevance for studying a wide range of diseases and included established risk factors for disease (e.g. lipids for vascular disease, sex hormones for cancer), diagnostic measures (e.g. HbA1c for diabetes and rheumatoid factor for arthritis) or markers of phenotypes that were not otherwise well assessed (e.g. renal and liver function).

### Data preparation for liver fat imputation

#### DXA

We downloaded whole-body DXA scans (n = 67,326; data field 20158) and abdominal MRI-derived PDFF measurements (n = 33,058; data field 40061) from the UKB^23^. PDFF measures were used as ground truth to train our ML algorithms for imputing liver fat content from DXA, metabolites and biomarkers.

#### Metabolites

We used metabolomics data from the first data release, comprising 249 metabolite measurements from a random selection of 118,461 baseline plasma samples in the UKB (data field 220). The metabolites dataset, downloaded from the UKB, underwent several processing steps. First, we removed fields that were not related to metabolite measurements, such as quality control flags. Next, we excluded all non-baseline measurements. Finally, we removed metabolites and samples with one or more missing values, resulting in a final dataset with 117,024 participants and 188 metabolites. We observed a strong pairwise phenotypic correlation between several metabolites (Supplementary Fig. 1). Therefore, we performed principal component analysis (PCA) on the metabolites dataset and used all principal components (PCs; same number as the number of metabolites) as inputs to our ML model (described below). We also included age, sex, and body mass index (BMI) terms - weight (in kilograms), inverse of height (in meters), and inverse of height squared - due to the potential for biased effect estimates from the inclusion of ratio covariates^29^, as inputs to our model.

#### Biomarkers

We selected all the biochemical markers measured in blood collected at baseline from all participants (data field 18518), and supplemented them with several hematological assay measures (blood counts, assayed at baseline) and anthropometry measures. We excluded markers with data missing from more than 10% of the participants. The final set of 37 measures - collectively referred to as biomarkers throughout this study - is shown in (Supplementary Table 1). With the inclusion of age and sex (as covariates), we ended up with a total of 39 traits. Most of these traits underwent a log transformation, except for those with a binary distribution (diabetes status and sex) or appearing normal (age).

### ML workflows for liver fat imputation

#### DXA-based imputation

Initially, we identified 28,908 subjects in the UKB with both whole-body DXA scans and abdominal MRI-derived PDFF estimates available. Subsequently, these samples were split into a training set (n = 23,126), a validation set (n = 2,890), and a test set (n = 2,892). During the study, the UKBB made DXA scans and PDFF estimates available for an additional 2,800 participants, constituting a second test set (test set 2).

We compared two deep learning architectures, ResNet-50^30^ and EfficientNet-B0^31^, for predicting PDFF and other adiposity terms, including visceral adipose tissue (VAT), abdominal subcutaneous adipose tissue (SAT), and gluteofemoral adipose tissue (GFAT) from DXA scans. The final layer of these models was replaced with a layer containing nodes corresponding to PDFF, VAT, SAT and GFAT. We used the ImageNet^32^ pre-trained models as input for the first epoch and fine-tuned the models using our training set. During the training process, we applied trimming to the whole-body DXA scans (removing 10% from the top and 25% from the bottom), rescaled them to 320x320, and then applied random data augmentation, including 10-degree rotations and 5% translations. These steps were implemented using PyTorch v2.0.1 and Torchvision v0.15.2.

#### Metabolite- and biomarker-based imputation

We first identified participants from both metabolite and biomarker modalities who also had PDFF measurements (n = 9,723 for metabolites; n = 40,533 for biomarkers). Within each modality, we implemented a 90:10 split, reserving 10% of the participants for model evaluation (the test set). In both modalities, we employed XGBoost^33^ regression using the input features described earlier (see data preparation) and log-transformed PDFF values as the outcome. Additionally, the time between the baseline and imaging visit was included as a feature in our models. For predictions on samples without an imaging visit, this term was set to zero. Removing this term did not significantly impact our model (data not shown). To optimize model performance, we conducted hyperparameter tuning using a five-fold cross-validation approach. We then retrained the models on the entire 90% split.

### Ablation analysis within the biomarker modality

The goal of the ablation analysis within the biomarker modality was to determine whether a single biomarker or a set of strongly correlated biomarkers predominantly influenced the model predictions. In order to identify markers that are strongly correlated with each other, we conducted a clustering analysis on the complete set of input features. Through visual inspection, we identified four sets of strongly correlated biomarkers (Supplementary Fig. 2). These sets were categorized as: i) overall body composition; ii) components of blood; iii) liver enzymes; and iv) lipid metabolism and cardiovascular health. These biomarkers were ablated as a set, while the remaining biomarkers were ablated individually (Supplementary Fig. 2 & Supplementary Table 2).

The ablation analysis model training followed a process similar to that of the full model; 10% of the data were set aside for evaluation, and hyperparameter tuning was carried out using five-fold cross-validation. The models were subsequently trained on the complete 90% split, and evaluation was performed on the reserved 10% of the data. To ensure consistency across ablated models, the same samples were used for both training and evaluation in each case, aligning with the approaches employed for the full model.

### Genotype data preparation

We downloaded the imputed genotype dataset release (version 3), and selected variants with minor-allele frequency > 0.001, imputation quality (INFO) score > 0.8, and Hardy-Weinberg disequilibrium *P* value > 1.0 x 10^-10^. Following Bycroft *et al*.^5^, we removed samples if they were flagged as outliers by missingness or heterozygosity, or displayed sex chromosome aneuploidy. Samples were further filtered down to unrelated samples with self-reported “White-British”, “White”, or “Irish” ancestry, and those that were within ±seven standard deviations of the first six genetic PCs^20^. The final dataset after quality control included 356,798 participants and 10,874,712 variants.

### Genome-wide association study (GWAS)

GWASs were conducted using an additive linear regression model with PLINK (v1.9)^34^ software. Abdominal MRI-derived PDFF (downloaded from the UKB) and liver fat percentages predicted from DXA, metabolites, and biomarkers were inverse normal transformed, and were regressed on genotypes using array type, age, age^2^, age^3^, sex, age*sex, age^2^*sex, age^3^*sex, top 20 genetic PCs, and BMI terms weight, 1/height and 1/height squared as covariates. We limited all our GWASs to unrelated participants of white British ancestry.

### Locus definition

We first resolved GWAS summary statistics to independent signals using PLINK’s^34^ linkage disequilibrium (LD) clumping (LD *r*^2^ threshold = 0.1 and window size = ±500 kb). Signals whose lead variants were separated by 1 Mb or less were subsequently merged into what we define here as a locus, retaining the variant with the strongest association. Any reference to loci henceforth refers to clumping with these parameters, unless otherwise noted.

### GWAS meta-analysis of clinical MASLD

We downloaded all four individual cohort-level, case-control GWAS summary statistics of clinically diagnosed MASL from Sveinbjornsson *et al*.^3^, and meta-analyzed them using the inverse-variance weighted average method based on a fixed-effects model implemented in META^35^. The meta-analyzed summary statistics included 9,491 patients with MASL and 876,210 population controls.

### Known loci

Based on the latest GWAS meta-analyses reported by Sveinbjornsson *et al*.^3^ and Chen *et al.*^4^, we identified 15 loci demonstrating a robust association with clinically diagnosed MASL, MASLD and/or PDFF as known loci. Briefly, Sveinbjornsson *et al*.^3^ conducted a GWAS on PDFF estimates (n = 36,116) in the UKB (our internal GWAS of PDFF included a subset of 23,658 of these individuals). Additionally, Sveinbjornsson *et al*.^3^ meta-analyzed GWAS summary statistics from four independent cohorts of clinically diagnosed MASL (identified using ICD9/10; n = 9,491 patients with MASL and 876,210 population controls). On the other hand, Chen *et al*.^4^, reported a meta-analysis of two independent cohorts of clinical MASLD and four independent cohorts of PDFF.

Of the two lead missense variants in *TM6SF2*, rs58542926 and rs187429064, reported by Sveinbjornsson *et al*.^3^, rs187429064 was removed as it was part of the same locus (see our locus definition above). Similarly, we omitted rs1229984, a missense variant in *ADH1B* that was reported in both the studies as it was only ∼250 kb away from the lead variant in *MTTP*. Additionally, rs17817449, an intronic variant in *FTO* and rs10756038, an intronic variant in *PTPRD* loci from Chen *et al*. were excluded because these loci primarily capture BMI, and our analysis was adjusted for BMI.

### SNP-based heritability and genetic correlation

We calculated SNP-based heritability of each trait and the genetic correlation (*r*_g_) between pairs of traits by conducting LD score (LDSC) regression^36,37^ analysis on the GWAS summary statistics. We used the precomputed LD scores based on European samples from the UKB as a reference (https://pan.ukbb.broadinstitute.org/docs/ld).

## Results

We leveraged ML to predict liver fat content using three different data modalities available in the UKB: DXA scans, plasma metabolites, and anthropometric and blood-based biochemical measures (biomarkers; Supplementary Table 1). We used abdominal MRI-derived PDFF measures available in the UKB as a ground truth. Within each modality (DXA, metabolites, or biomarkers), we partitioned the dataset to create at least one hold-out independent test set, ensuring it was not used during the model training or tuning. We used the R^2^ statistic on these test sets as the primary measure to assess our model performance. The imputed liver fat percentages were then used to conduct GWASs, separately within each modality, alongside MRI-derived PDFF. An overview of our study is presented in Fig. 1.

**Fig. 1:**
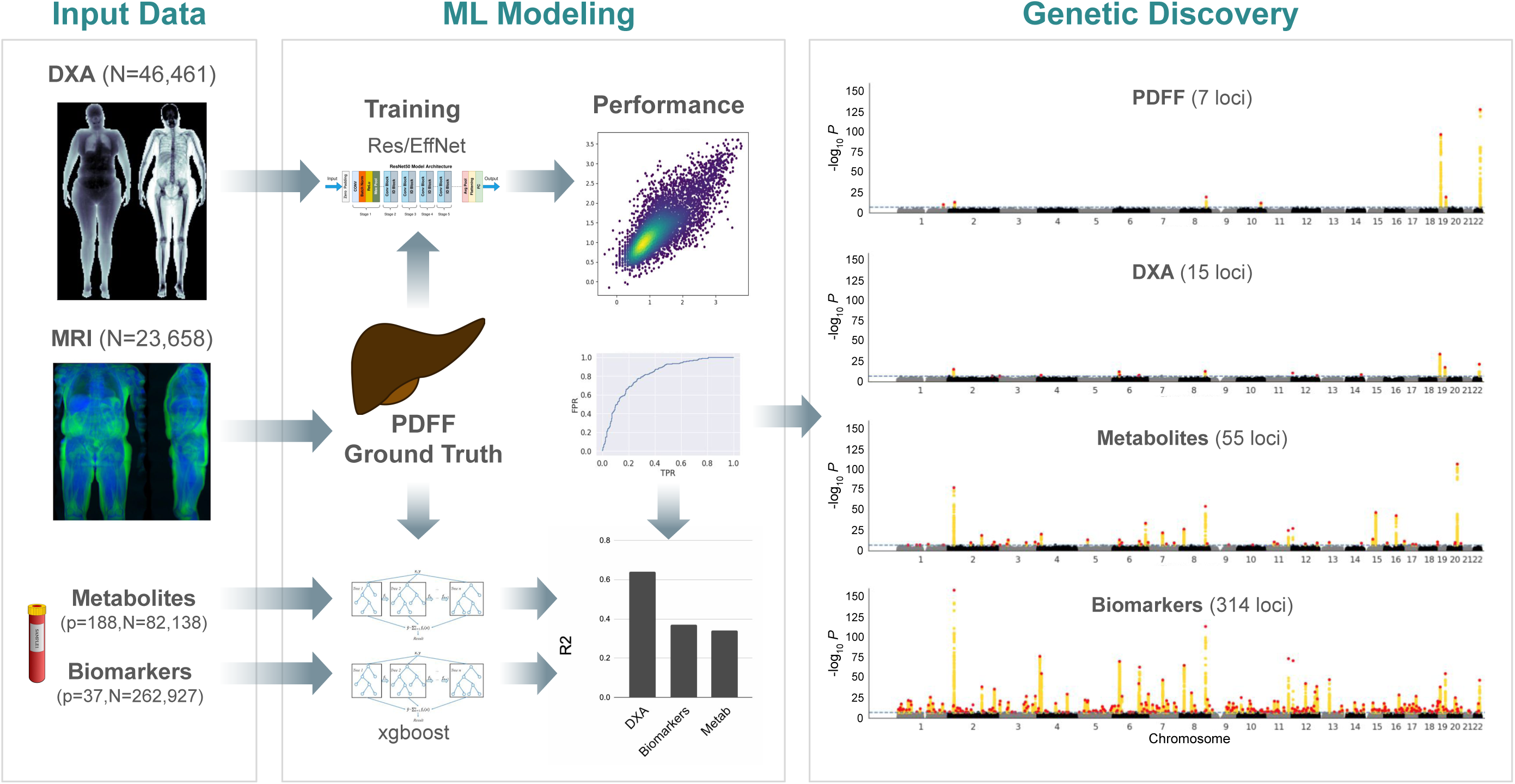
Study overview. ***Input Data***: We imputed liver fat content using three distinct data modalities, DXA, metabolites and biomarkers, in the UKB. We used abdominal MRI-derived PDFF as ground truth. ***ML Modeling***: We used two deep learning architectures, ResNet-50 and EfficientNet-B0 to predict liver fat from DXA, and XGBoost models to predict from both metabolites and biomarkers. We used R^2^ and AUC-ROC in the test set to assess our model’s performance. ***Genetic Discovery***: We then performed GWASs on both PDFF and imputed liver fat percentages.

### Liver fat prediction from DXA, metabolites, and biomarkers

#### DXA scans

Following the comparison of fine-tuning using either EfficientNet-B0 or ResNet-50 on the training and validation sets, we observed superior performance with EfficientNet-B0, achieving R^2^ values of 0.63 and 0.65 in test set 1 and test set 2 (see Methods), respectively (Fig. 2). These results indicate that our models could explain approximately 63-65% of the variance in abdominal MRI-derived PDFF, reflecting a substantial correlation between the predicted and actual values (rho = 0.80; Fig. 2). To further assess the utility of these predictions, we evaluated their ability to predict MASLD case-control status, defined by the PDFF threshold of 5.5% in the test set. We observed an area under the receiver operating characteristic (AUC-ROC) curve value of 0.90 (Fig. 2).

**Fig. 2:**
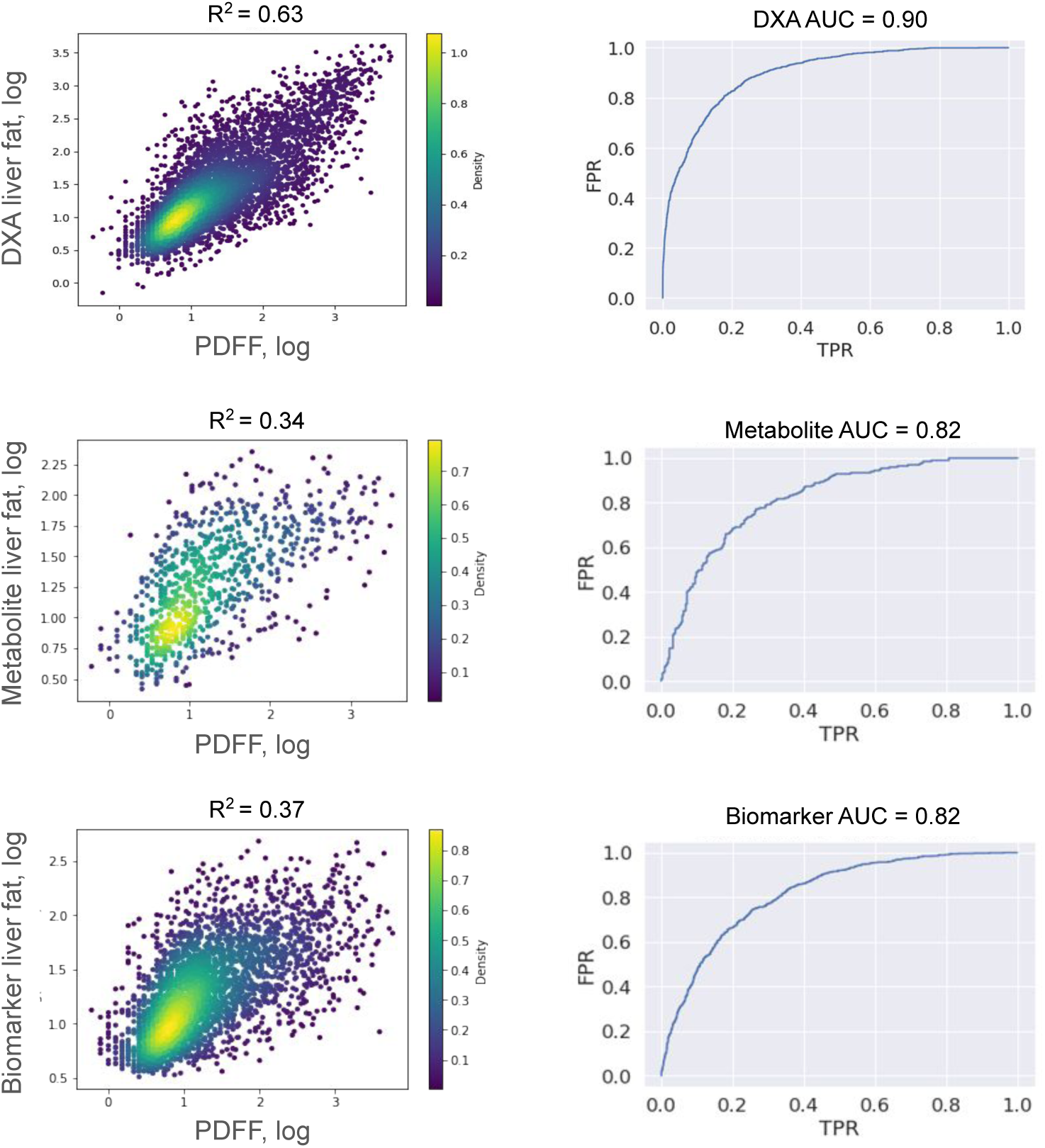
Performance evaluation of the three models using the PDFF truth set. ***Left***: Scatter plots illustrating the relationship between abdominal MRI-derived PDFF (ground truth; *x* axis) and ML-imputed liver fat content (*y* axis) from DXA, metabolites and biomarkers. ***Right***: AUC-ROC curves demonstrating the ability of each of the three models to predict MASLD case-control status, defined based on a PDFF threshold of 5.5%, in the test set is shown on the right. The top panel represents data from DXA, the middle panel from metabolites, and the bottom panel from biomarkers

To further interpret the DXA model, we conducted two follow-up analyses. First, we manually inspected gradient-based saliency maps to assess the importance of different regions in the input whole-body images for the model’s prediction. Examples in Supplementary Fig. 3 show that the area around the liver exhibits meaningful classification gradients in two individuals. Notably, the relative saliency of the liver is more pronounced in leaner subjects compared to individuals with greater peripheral adiposity, possibly reflecting an enhanced ability of the model to assess liver fat content in leaner individuals. Second, we performed masking and subsetting studies to systematically illustrate the importance of the liver area to our model. We masked the left or right half of the images in the training and validation sets, reran the training process, and tested the prediction performance on the validation set. An example of masking is shown in Supplementary Fig. 3. Additionally, we tested subsetting the images into one of four quadrants, limiting input data to one quadrant at a time for liver fat predictions. This approach enabled a nuanced evaluation of the model’s sensitivity to different regions of the images (Supplementary Fig. 3).

The results of these studies are summarized in Supplementary Table 3. Taken together, these results demonstrate that higher performance liver fat prediction is achieved only when the liver area is retained in the images, corresponding to the left side in the left-right experiment, and the top-left quadrant in the quadrant-based experiment. In contrast, when the liver area is absent in the input data, validation set R^2^ scores drop from 0.58 or greater to less than 0.34, representing a >40% decrease in the predictive performance.

#### Metabolites and biomarkers

While the test set R^2^ values for metabolite-based (R^2^ = 0.34) and biomarker-based (R^2^ = 0.37) predictions were notably lower than those obtained from DXA, scatter plots revealed a linear trend between PDFF measurements and predicted liver fat percentages for both modalities (Fig. 2). To further investigate the practical utility of these predictions, we assessed their ability to predict MASLD cases from controls, defined by the PDFF threshold of 5.5% in the test set. Notably, both metabolites and biomarkers exhibited high AUC-ROC values (0.82 for both), suggesting their effectiveness in predicting MASLD status (Fig. 2).

Next, to explore whether individual biomarkers might be disproportionately influencing our predictions, we conducted an ablation study. These results revealed an extremely strong correlation with liver fat predicted from the full model, and the set R^2^ showed no meaningful change regardless of the ablation set used (Supplementary Fig. 4). This observation persisted even when removing single biomarkers or the cluster of strongly correlated markers (Supplementary Fig. 4).

### MASL is largely underdiagnosed in the UKB

While 848 (2%), 2,133 (3%) and 5,575 (2%) of the UKB participants with DXA, metabolites and biomarker-imputed liver fat content received an ICD9/10 diagnosis code for MASL (k76), 11,565 (25%), 20,225 (25%) and 75,653 (29%) met the criteria for MASL, defined as liver fat content >5.5% (Table 1). In alignment with our ML-based imputations, 5,725 (24%) participants with MRI-derived PDFF measurements met the criteria for MASL, while only 430 (2%) of the 23,658 individuals have received a clinical diagnosis (Table 1). These findings are consistent with a previous report^7^.

### Genetic associations of liver fat

We conducted GWASs, separately, for MRI-derived PDFF (n = 23,658), liver fat percentage predicted from DXA (n = 46,461), metabolites (n = 82,138) and biomarkers (n = 262,927), restricting to unrelated individuals of white British ancestry (see Methods). In line with the effective sample size, our GWAS revealed seven loci associated with PDFF (*h*^2^_SNP_ = 0.18, se = 0.036), 15 with DXA (*h*^2^_SNP_ = 0.17, se = 0.015), 55 with metabolites (*h*^2^_SNP_ = 0.19, se = 0.02), and 314 with biomarker-derived liver fat (*h*^2^_SNP_ = 0.23, se = 0.015; Fig. 3). We observed inflation of the summary statistics (lambda GC = 1.09, 1.26, and 1.73 for DXA, metabolites, and biomarkers, respectively); however, the LD score regression (LDSC) intercept (1.02, 1.08, and 1.12 for DXA, metabolites, and biomarkers, respectively) and the ratio (0.13, 0.20 and 0.09 for DXA, metabolites and biomarkers, respectively) suggests that these associations are primarily driven by polygenicity. Of the seven PDFF-associated loci in our study (Table 2): 5, 2, and 5 respectively were replicated by DXA, metabolites, and biomarkers at genome-wide significance; 7, 5, and 7 had effects in the same direction; and 6, 2, and 7 of these directionally consistent loci showed at least nominal evidence of replication (*P* < 0.05; Table 2).

**Fig. 3:**
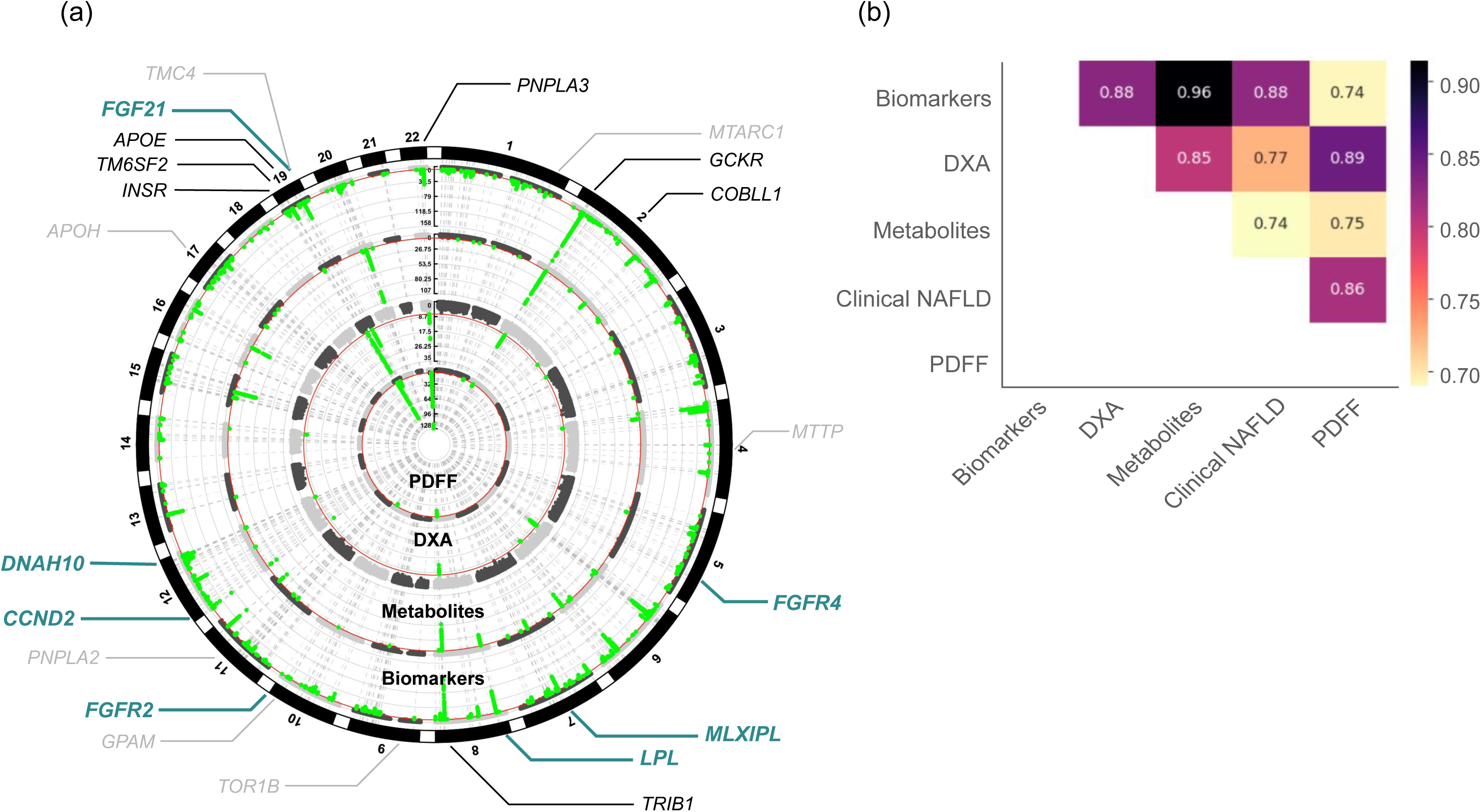
GWAS associations of ML-imputed liver fat and PDFF. (a) Circos plot showing four bands (from center to periphery) corresponding to -log_10_ *P* value of association of genetic variants with abdominal MRI-derived PDFF, and liver fat predicted from DXA, metabolite, and biomarkers. The red line indicates genome-wide significance (*P* = 5 x 10^-8^). Black gene labels represent known loci replicated at genome-wide significance in one or more of our modalites. Gray gene labels indicate known loci that did not get replicated at genome-wide significance. Teal gene labels denote some of the newly identified loci. (b) Heatmap demonstrating the pairwise genetic correlation estimates (*r*_g_) among liver fat derived from across our modalities, abdominal MRI-derived PDFF and clinical MASLD.

### Genetic correlations across modalities

To assess the extent to which genetic variation explains the correlation between traits, we estimated the genetic correlation between liver fat derived from DXA, metabolites, biomarkers, PDFF, and clinically diagnosed MASLD^3^ (see Methods). Because the GWAS from clinical MASLD was not adjusted for BMI, we used liver fat summary statistics that were similarly not adjusted for BMI from all three data modalities, alongside MRI-derived PDFF. We found a strong genetic correlation between all pairs of traits with (*r*_g_ ranging from 0.74 to 0.89, with the strongest correlation of 0.89 observed between DXA and PDFF; Fig. 3b). Overall, these results suggest a substantial shared genetic basis for liver fat predicted from the three modalities, and supports the relevance of our ML-imputed liver fat genetic associations to both PDFF and, more importantly, clinically diagnosed MASLD. Notably, biomarker-derived liver fat demonstrated a genetic correlation of 0.88 with clinical MASLD, suggesting that a majority of the newly-identified loci are likely to be clinically relevant (Fig. 3b).

### Relative value of each modality for genetic discovery in MASLD

To assess the effectiveness of each of the three data modalities for genetic discovery in MASLD, we first evaluated the ability of our ML-imputed liver fat to detect known associations. Considering variations in sample size across modalities, we employed directional consistency assessments along with nominal evidence of association as a metric. Among the 14 of the 15 previously known loci (see Methods) present in our study, 14, 10 and 13 exhibited a directionally consistent effect on liver fat derived from DXA, metabolites and biomarkers, respectively, and 12, 5 and 13 of these showed an evidence of replication at *P* < 0.05 (Table 2). While certain known associations, such as *GCKR*, *COBLL1*, *TRIB1* and *INSR*, demonstrated an enhanced strength of association with liver fat in line with increased sample size across modalities from DXA to biomarkers irrespective of the prediction accuracy, others, including *MTARC1*, *GPAM*, *TMC4*, *TM6SF2* and *PNPLA3*, exhibited a diminished strength of association (Table 2 & Supplementary Fig. 5). This suggests that ML imputed liver fat predictions capture multiple mechanistic components of MASLD, with some components being more precisely captured while others appear to be noisier. Surprisingly, the association between liver fat predicted from metabolites and the two well known MASLD loci, *APOE* and *PNPLA3*, was directionally inconsistent, suggesting a potential modality-specific nature of our liver fat predictions in capturing certain pathobiological or mechanistic components of MASLD (Table 2).

Next, we employed precision-recall analysis to evaluate the performance of our modalities (Supplementary Fig. 6). We defined precision as the fraction of ML-imputed liver fat loci in our study that were among the known loci, and recall as the fraction of known loci that were detected by our imputed liver fat GWASs. We observed that while DXA modality was most informative (67% precision and 75% recall), plasma metabolites were least informative (18% precision and 50% recall). Lastly, liver fat predicted from biomarkers yielded an 88% recall value, but only had a 15% precision. However, the observed lower precision for biomarkers could be attributed to limited statistical power in clinically diagnosed MASLD case-control GWASs, potentially hindering the identification in those studies of many of our biomarker-derived liver fat associations.

### Biomarker-predicted liver fat loci are less likely to be driven by individual markers

To assess whether our biomarker-derived liver fat loci were driven by one or more of the individual biomarkers used as inputs to our model as opposed to capturing the “aggregate trait” (liver fat), a series of GWASs were conducted for liver fat values predicted using leave-one-out approach. Because we noticed an extremely strong phenotypic correlation between some of the markers, resulting in clusters, we also predicted liver fat using the leave-one-cluster-out approach (Supplementary Fig. 2 & Supplementary Table 2). In line with phenotypic correlations and prediction accuracies, (Supplementary Fig. 4), genetic correlations at the level of the whole genome yielded similar results when compared with the main analysis using liver fat predicted from all the biomarkers (Supplementary Fig. 7).

### Expanding the genetic landscape of liver fat accumulation and MASLD

Considering the remarkably strong genetic correlation with PDFF and clinically-diagnosed MASLD, we next evaluated the utility of our ML-imputed liver fat GWASs to further expand our understanding of the biology of clinical MASLD. Across DXA, metabolites and biomarker modalities, we identified 15, 55 and 314 loci, respectively, totalling 321 independent loci collectively; of these, 10, 51, and 305 are novel contributing to a total of 312 novel loci (Fig. 3).

Next, of the 10, 51 and 305 of our newly identified loci from DXA, metabolites and biomarkers, respectively, 6, 27 and 196 had data available in the clinically diagnosed MASLD case-control GWAS summary statistics^17^; of these, 5 (83%), 18 (67%) and 153 (78%) showed directionally concordant effects on clinical MASLD (Supplementary Fig. 8). We also compared our findings to those identified by Miao *et al*., from the GWAS of their biomarker-imputed MASLD case-control status in the UKB, observing a strong correlation between the effect estimates (Supplementary Fig. 8). Collectively, these findings suggest that as the sample size of clinical MASLD GWAS (the number of cases in particular) increases, many of our newly identified ML-imputed liver fat associations are likely to become more apparent.

Among the newly identified associations are *FGF21* and its receptors *FGFR2* and *FGFR4*, *MLXIPL* and *LPL*. There are multiple clinical trials prosecuting FGF21 agonism as a therapeutic strategy against the whole spectrum of MASLD, including steatosis, fibrosis and cirrhosis^38,39^. However, to our knowledge, there was no prior human genetics support for these targets.

The *MLXIPL* gene encodes a transcription factor carbohydrate response element-binding protein (ChREBP) that is involved in the regulation of various genes, including *FGF21* - one of our novel associations - and *GCKR* - a well known locus for MASLD, related to carbohydrate and lipid metabolism. In the context of MASLD, *MLXIPL* is recognized as a master regulator of *de novo* lipogenesis, a process by which the liver synthesizes fatty acids from non-lipid precursors, such as carbohydrates^40^. Increased *de novo* lipogenesis is often observed in individuals with MASLD, contributing to the accumulation of fat in the liver^40,41^. In addition, expression of *MLXIPL* has been reported to be altered in individuals with MASLD^40^.

LPL is an enzyme involved in lipid metabolism, playing a crucial role in the hydrolysis of triglycerides, which can affect the availability of fatty acids for storage and energy production. In relevance to MASLD, Ghodsian *et al.*^42^ demonstrated that genetically predicted *LPL* expression in subcutaneous adipose tissue is associated with clinically diagnosed MASLD using transcriptome-wide association analysis. Furthermore, using a Mendelian randomization framework, Li *et al.*^43^ identified that among nine lipid-lowering drug targets, LPL is the only drug target associated with a lower MASLD risk.

### Liver fat associations with multimodal support

Next, we further evaluated liver fat content associated loci with concurrent support from multiple modalities. Having GWAS loci identified in more than one data modality offers several advantages. First, it mitigates modality-specific biases, providing a form of cross-validation that enhances confidence in the robustness and reproducibility of the associations. Second, it strengthens the argument for the biological relevance of the identified loci. A majority of our loci with genome-wide significant association in at least one modality demonstrated replicative evidence in one or both of the remaining modalities. The breakdown of all the 321 independent loci (collectively identified from across our three modalities) and their varying level of multimodal support at *P* < 0.05 / 321 is presented in Fig. 4a.

**Fig. 4:**
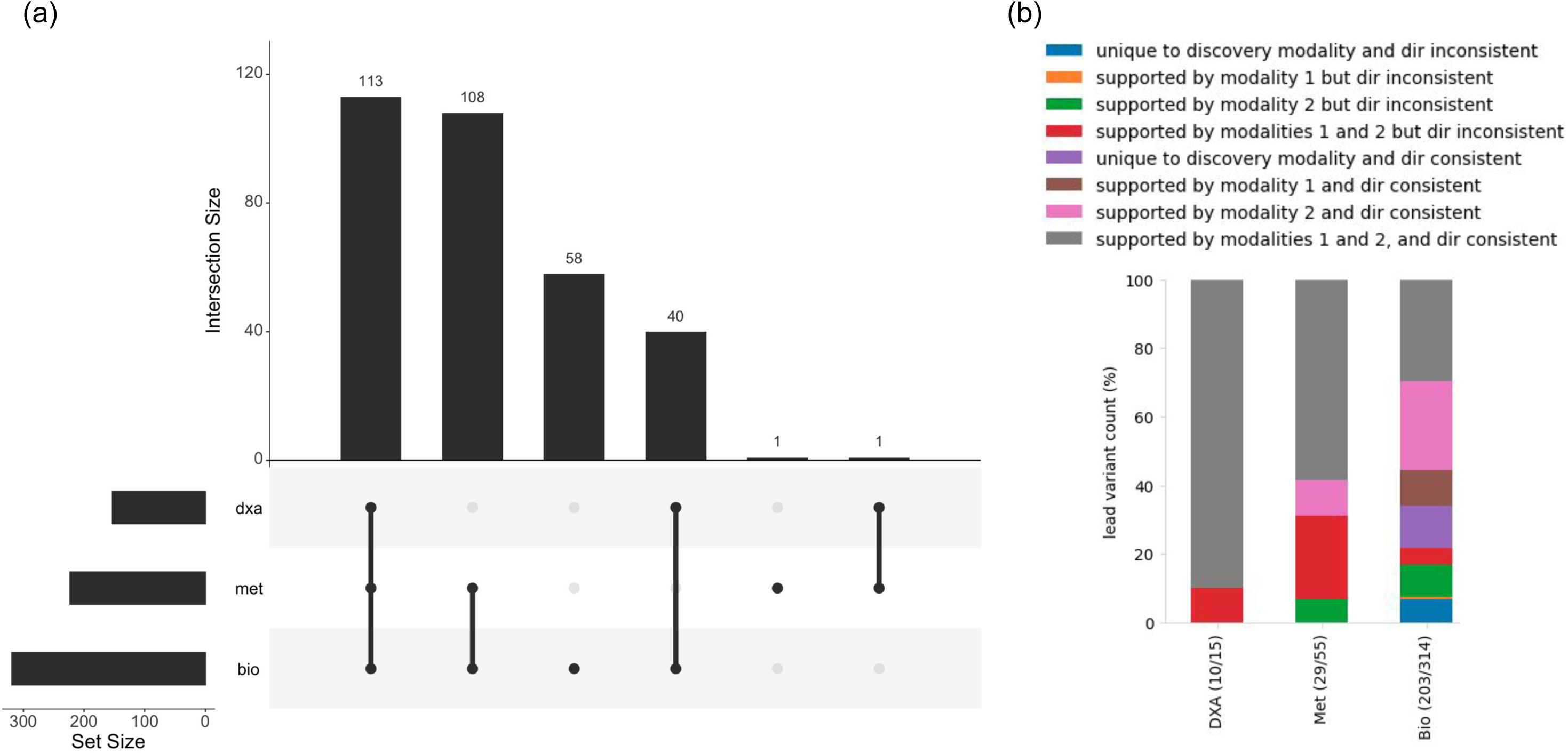
Loci with multimodal support and their likely relevance to clinical MASLD. (**a**) UpSet plot depicting the counts of the total number (from across all 3 of our modalities) of genome-wide significant independent loci and their breakdown by multimodal support. (**b**) Histogram depicting the proportion of loci within each modality, demonstrating varying degrees of multimodal support, and their directionally consistent effects on clinically diagnosed MASLD. Only loci with data available in clinical MASLD summary statistics are considered (10, 29 and 203 from DXA, metabolites and biomarkers, respectively). When DXA is used as the discovery modality, metabolites are considered as modality 1 and biomarkers as modality 2. For metabolites, DXA and biomarkers are considered as modalities 1 and 2, respectively. For biomarkers, DXA and metabolites are considered as modalities 1 and 2.

Given the varying prediction accuracy and sample size across modalities - factors that could contribute to differences in statistical power - we evaluated multimodal support for each locus with genome-wide significant association in the discovery modality by utilizing discovery modality-specific Bonferroni-corrected *P* value from the other modalities as replicative evidence (*P* < 0.05 / the number of loci within the discovery modality). First, within each modality, we categorized the genome-wide significant loci into four groups based on the level of multimodal support (unique to discovery modality, replicated by modality 1, modality 2, or both), and compared the mean GWAS *P* values across these groups. We observed that, as the number of modalities with replicative support increases, the mean *P* value of association in the discovery modality becomes stronger (Supplementary Table 4). For example, the mean (-log_10_) *P* value for biomarker-derived liver fat loci with support from both DXA and metabolite modalites was 20.70 (median *P* = 1.2 x 10^-14^; group size = 110 loci). In comparison, the mean values were 17.70 (median *P* = 2.0 x 10^-12^; 150 loci) for loci with support from DXA only, 17.25 (median *P* = 7.8 x 10^-13^; 217 loci) for those supported by metabolites only, and 10.93 (median *P* = 8.6 x 10^-10^; 57 loci) for loci unique to biomarkers.

Next, we evaluated whether loci with multimodal support are more likely to be relevant for clinical MASLD by assessing the proportion of loci with varying degrees of multimodal support that demonstrated a directionally consistent effect on clinical MASLD. For instance, out of the 314 biomarker-derived liver fat loci (where we have the most loci), data were available for 203 in the clinical MASLD GWAS summary statistics. Among these, 92, 142 and 70 had supporting evidence from DXA, metabolites, and both DXA and metabolites, respectively (Fig. 4b). Of these, 81 (88%), 113 (80%) and 60 (86%) demonstrated a directionally consistent effect on clinical MASLD while only 64% (25 of the 39 for which data was available in clinical MASLD GWAS) of the loci that were unique to biomarkers showed a directionally concordant effect (Fig. 4b). Taken together, these results are consistent with the idea that loci with multimodal support are more likely to be robust and biologically relevant.

## Discussion

The advent of large-scale biobanks, spearheaded by initiatives like the UKB, has unlocked numerous opportunities for genomics-based target discovery. In this study, we expanded the genetic catalog of liver fat accumulation significantly, utilizing both deep (small sample size with a high-resolution phenotype, e.g. DXA) and shallow (large sample size with low-resolution phenotype, e.g. biomarkers) phenotypes, both imputed using ML. We identified a total of 321 independent loci associated with liver fat content, which is 45 times more loci than previously reported. More importantly, our findings demonstrate that many of these newly identified loci are likely clinically relevant to MASLD.

Given the extent of underdiagnosis in MASLD, the quantification of liver fat has emerged as a crucial metric for assessing disease status. Imaging modalities, particularly MRI, have proven effective not only in quantifying liver fat, but also revealing its genetic underpinnings. Both traditional and ML-based MRI processing approaches have contributed to our understanding of the genetic basis of liver fat accumulation^3,7,10,44^. However, relying solely on MRI as the primary modality presents challenges, especially in large-scale biobanks like the UKB, where the number of participants with MRI scans is limited. To overcome this limitation, our study leveraged ML frameworks across a comprehensive dataset, integrating results from various modalities to further expand our understanding of the genetic basis of liver fat accumulation.

Our ability to predict liver fat content from DXA with high accuracy is a considerable improvement over prior studies. For instance, Bazzocchi *et al.* manually identified six liver-associated regions of interest in DXA scans from 90 patients with hepatic steatosis and 90 healthy controls, and demonstrated the utility of DXA in evaluating hepatic steatosis, achieving AUC of 0.82^14^. In another study, Bouchi *et al.* compared the DXA-derived android-to-gynoid (A/G) ratio to liver attenuation index (LAI) assessed by abdominal computed tomography in a small cohort of 259 patients with diabetes, observing a significant association^15^. More recently, Tan *et al.* investigated MASLD risk in 10,865 non-obese and 16,487 obese individuals using DXA-derived regional fat percentage scores^16^. Their multivariate logistic regression analysis, accounting for age, race, BMI, and diabetic status, revealed increased MASLD risk with regional fat percentage in both obese and non-obese subjects. While these studies demonstrated significant statistical correlations between DXA-derived features and MASLD or MASLD-related traits, they are limited by factors such as small sample size, reporting performance on training sets without independent test sets, and the absence of reliable ground-truth (continuous) measures, such as PDFF. The prediction performance we obtained using deep learning models on much greater sample sizes, both for MASLD case-control status (AUC = 0.90) and especially for liver fat percentage (R^2^ = 0.64) represent a major improvement, and suggests a potential for using DXA in clinical care or clinical trials to predict at-risk patients for treatment. Furthermore, this study represents the first demonstration of using DXA-derived liver fat to recapitulate and to further expand the genetic landscape of MRI-derived PDFF or clinically diagnosed MASLD.

While the phenotypic accuracy of liver fat predicted from biomarkers and metabolites was modest, we hypothesized that an increased sample size could mitigate the noise in our estimates. Supporting our hypothesis, these modalities - particularly biomarkers - not only replicated a majority of known loci but also enabled us to significantly expand the genetic landscape of liver fat accumulation. This highlights the potential of an ML-derived “shallow” phenotype, at sufficient sample size, to identify meaningful genetic associations. While no prior studies attempted to predict liver fat from biomarkers, Miao *et al*.^17^ used a subset of these markers (n = 14), including liver enzymes (ALT, AST, AST/ALT and GGT), lipids (triglycerides, cholesterol), diabetes-related traits (HbA1C, albumin, type 2 diabetes status), anthropometric measures (waist circumference, BMI), and covariates (age, age^2^, sex) and created a synthetic MASLD case-control cohort in the UKB to enable a GWAS, ultimately yielding 94 loci. We noticed a strong correlation in effect estimates between the loci identified by Miao *et al*., for biomarker-imputed MASLD status and our liver fat GWASs, particularly within the biomarker modality (see Supplementary Fig. 8). Because the genome-wide summary statistics from Miao *et al*. were not publicly available, we could not compare the effect estimates of all the loci that we identified. Nonetheless, the increase in the number of discovered loci in our ML-imputed liver fat GWAS compared to the recent ML-based GWAS by Miao *et al*. is consistent with increased sample size and/or the utilization of a quantitative trait instead of binary labels (GWAS of 28,396 ML-imputed MASLD cases and 108,652 population controls versus GWAS of ML-imputed liver fat in 262,927 participants).

To our knowledge, there have been no prior attempts to predict liver fat from metabolites at this scale, and their potential for genetic discovery remains unexplored. In contrast to the relatively strong performance observed in DXA and biomarker modalities, metabolite-predicted liver fat exhibited limited precision and recall, particularly in identifying known MASLD genetic associations. Noteworthy here is the inconsistency in the detected direction of effect at well-known loci such as *APOE* and *PNPLA3*. However, these limitations could be attributed to the combination of modest sample size, overall suboptimal prediction accuracy, and the potential presence of modality-specific biases in capturing certain mechanistic components of MASLD. Nonetheless, metabolite-derived liver fat replicated many genetic loci identified in other modalities and demonstrated a strong genome-wide genetic correlation with other modalities and clinical MASLD.

The strong genetic correlation between liver fat predicted across modalities and clinical MASLD suggests that our ML-imputed liver fat captures similar disease genetics. This implies that a majority of the loci associated with our ML-based predictions are highly likely to be relevant to the clinical disease of interest. Additionally, the high concordance in the genetic architecture of liver fat accumulation across modalities supports the contention that several genetic discoveries can be replicated across modalities, bolstering our confidence in these associations as it would diminish any potential modality-specific biases that may exist.

To our knowledge, this is the first deliberate use of a multimodal approach, leveraging both imaging (deep phenotype) and blood biochemistry (shallow phenotype) data, for the large-scale prediction of liver fat content. This strategy not only robustly underscored the extent of underdiagnosis of MASLD in the general population (such as the UKB) but also enabled the largest GWAS of quantitative hepatic fat. Additionally, our multimodal approach successfully demonstrated that ML-based phenotyping shows promise for improving both phenotypic accuracy, as evidenced in our DXA-based findings, and scalability, as observed in our biomarker-based findings. Our DXA findings, in particular, underscore the potential of employing deep learning techniques on more widely available, non-invasive imaging modalities for assessing hepatic steatosis. This reinforces the claim that imaging modalities often contain rich and complementary information that is biologically relevant.

The 312 novel liver fat-associated loci discovered by ML-based phenotyping substantially expand our knowledge of the underlying mechanistic processes of liver fat accumulation and clinical MASLD. Of particular note were liver-fat associated variants in/near genes involved in *de novo* lipogenesis (including *GCKR*, *TRIB1*, *INSR* and *MLXIPL*), one of the very well characterized mechanistic causes of MASLD. Furthermore, our data offer the first human genetic support for the role of FGF21 and its receptors in MASLD. Others have demonstrated increased clinical success for therapeutic agents with human genetic support^45–47^. In light of the positive findings of FGF21 in the clinical settings^39^, it was encouraging to see this association become evident and likely lending additional credence to this therapeutic strategy.

Lastly, the utilization of data from diverse modalities not only mitigates modality-specific biases but also provides a valuable form of cross-validation, enhancing confidence in the reproducibility of the associations. Our examination of liver fat associations with multimodal support underscores the robustness and biological relevance of the identified loci. The categorization of loci from each discovery modality based on the level of support from other modalities revealed a positive correlation between the number of supporting modalities and the strength of association in the discovery modality. This observation strengthens the validity of our findings and suggests that loci with multimodal support are more likely to be biologically significant. Furthermore, our assessment of clinical relevance demonstrated that loci supported by multiple modalities exhibit a higher likelihood of consistent effects on clinical MASLD. This aligns with the notion that these loci, validated across diverse data sources, are more robust and potentially crucial in the context of liver fat accumulation and clinically diagnosed MASLD. Overall, our findings provide valuable insights into the biological underpinnings of liver fat content and its clinical implications.

Our study has certain limitations. First, alterations in certain biomarkers we considered may be a consequence of hepatic steatosis rather than falling on the causal path, and some of the genetic associations may be driven by these markers. However, our sensitivity analysis suggests that our ML predictions capture the “aggregate trait” (liver fat) and are less likely to be driven by any individual marker or a cluster of strongly correlated markers. Second, the variability in prediction accuracy and sample size across modalities presents a challenge for conducting cross-modality comparisons. Nevertheless, multiple lines of evidence indicate that our ML-based liver fat predictions show promise for genomic discovery in MASLD, including the differentiation of MASLD subjects (PDFF measurements >5.5%) from non-MASLD controls, the replication of known genetic loci, the identification of plausible novel genetic associations, and the demonstration of strong shared genetic basis with clinically diagnosed MASLD. In summary, our results highlight the value of incorporating multimodal approaches in ML-enabled genetic studies, and demonstrate that orthogonal modalities can shed new light on the mechanistic and pathobiological underpinning of MASLD. This approach could be extended to other conditions and could be used to drive both new discoveries and to improve diagnostics.

## Tables and Supplementary tables

Table 1: Diagnosis rate based on ICD10 (k76) versus imaging/biomarker based criteria

Table 2: Known loci and their sumstats from our modalities

Supplementary Table 1: Final list of biomarkers used as input features to predict liver fat in the biomarker modality.

Supplementary Table 2: Individual biomarkers or groups removed from the input feature set during our ablation study

Supplementary Table 3: Results of the left-right masking, and quadrant-based subsetting studies within the DXA modality

Supplementary Table 4: Mean P value of loci with varying degrees of multimodal support

## URLs

- UK Biobank data fields: PDFF measures from data field 40061, DEXA from data field 40061, biomarkers from data category 18518, metabolites from data category 130

- GWAS implemented in Redun: https://github.com/insitro/redun

## Data availability

Summary statistics for GWAS presented in this study will be made available upon publication.

## Acknowledgements

The authors would like thank the participants of the UK Biobank, whose data were used with permission. This research was conducted using the UK Biobank Resource under approved Application Number 51766.

## Author Contributions

HS, SM, DA, DK, DJL, CO conceived of the project. HS, SM, DA, JP, KG, TS, CO performed analyses. All authors provided scientific input. All authors contributed to critical revision and/or writing of the final manuscript.

## insitro Research Team Banner and Contribution Statements

All contributors are listed in alphabetical order.

Downloading, preprocessing, and curation of UK Biobank data: Francesco Paolo Casale,¹,²,³,lJ Eilon Sharon,¹ Thomas W. Soare,¹ Baris Ungun^5^, James Warren¹

Software engineering: *Statistical genetics pipeline and bioinformatics resources:* Francesco Paolo Casale,¹,²,³,lJ Liz Ellithorpe,¹ Anna Merkoulovitch,¹ Colm O’Dushlaine,¹ Anna Shcherbina,¹ Thomas W. Soare,¹ Paul Sud,¹ Simon Tucker,¹ Baris Ungun¹; *redun development:* Robin Betz,¹ Edward Chee,¹ Patrick R. Conrad,¹ Kevin Ford,¹ Christoph Klein,¹ Donald Naegely,¹ Matthew Rasmussen¹

Affiliations: ¹insitro, South San Francisco, CA, USA; ^2^Institute of AI for Health, Helmholtz Munich, Neuherberg, Germany; ^3^Helmholtz Pioneer Campus, Helmholtz, Munich, Neuherberg, Germany; ^4^School of Computation, Information and Technology, Technical University of Munich, Munich, Germany; ^5^Therapanacea, 7 bis Bd Bourdon, 75004 Paris, France

---

**Supplementary Fig. 1:**
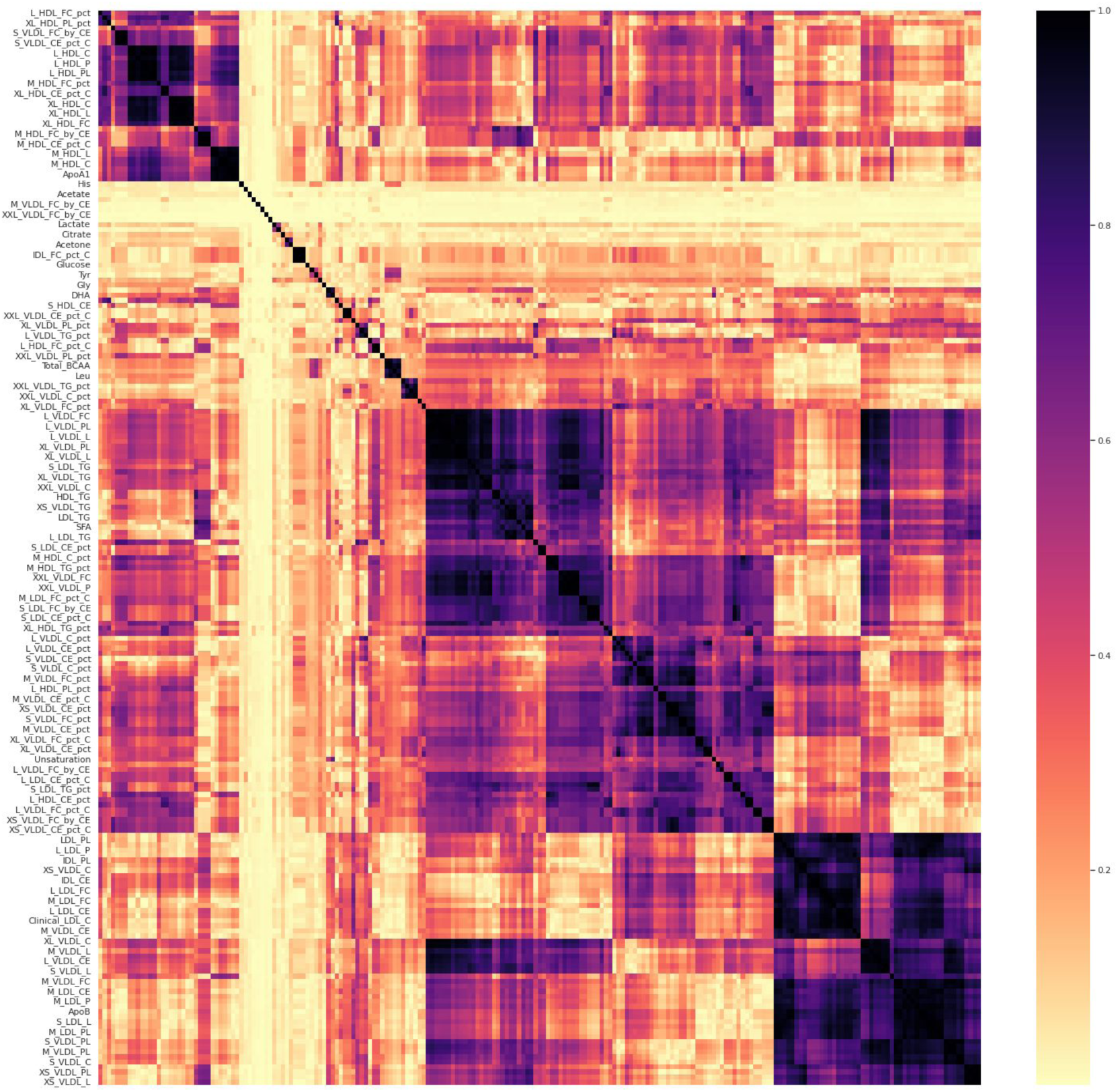
Correlation between metabolites. Heatmap demonstrating the pairwise phenotypic correlation between the metabolites used as input features. Color coded based on the absolute value of Spearman correlation

**Supplementary Fig. 2:**
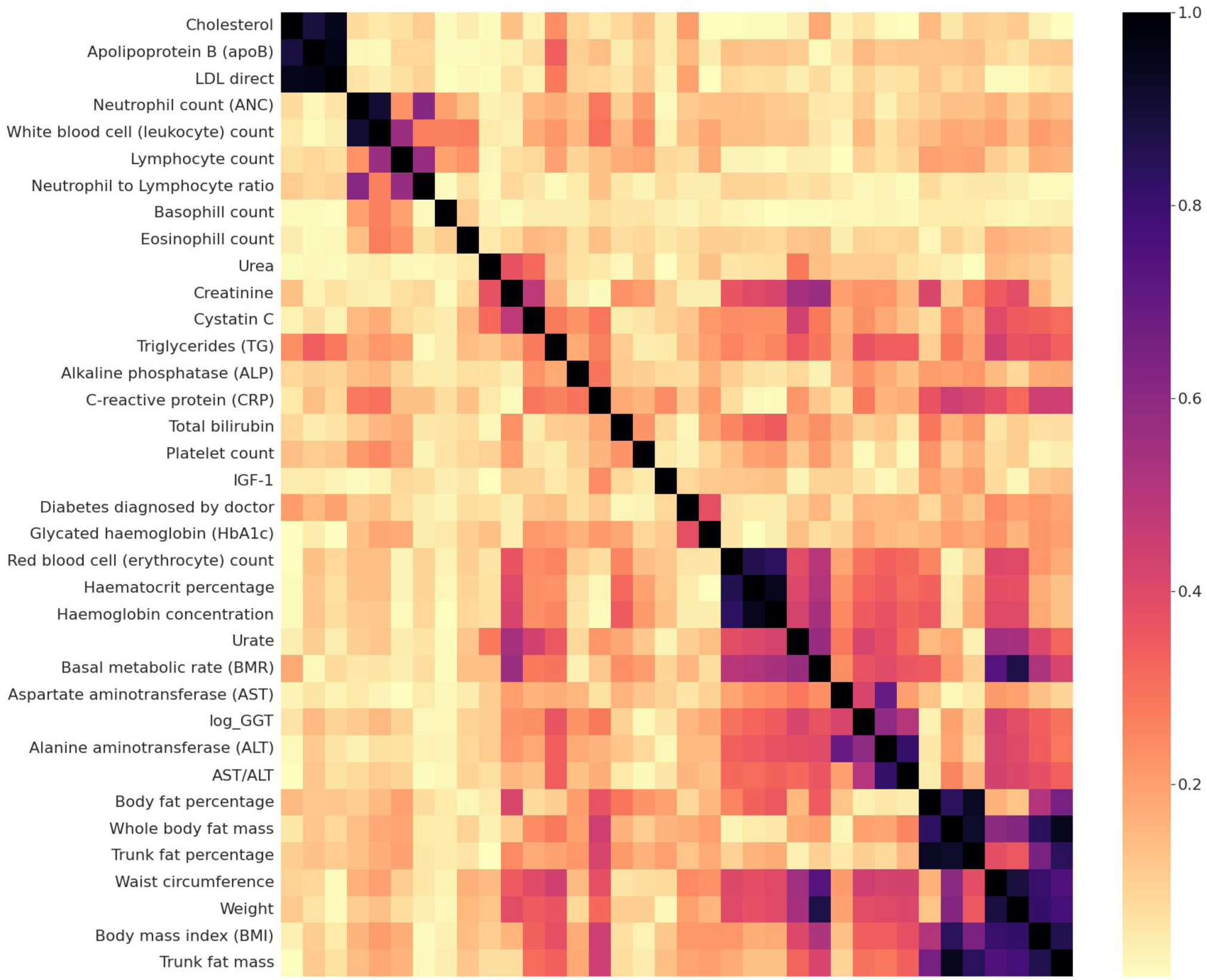
Correlation between biomarkers. Heatmap demonstrating the pairwise phenotypic correlation between the biomarkers used as input features. Color coded based on the absolute value of Spearman correlation

**Supplementary Fig. 3:**
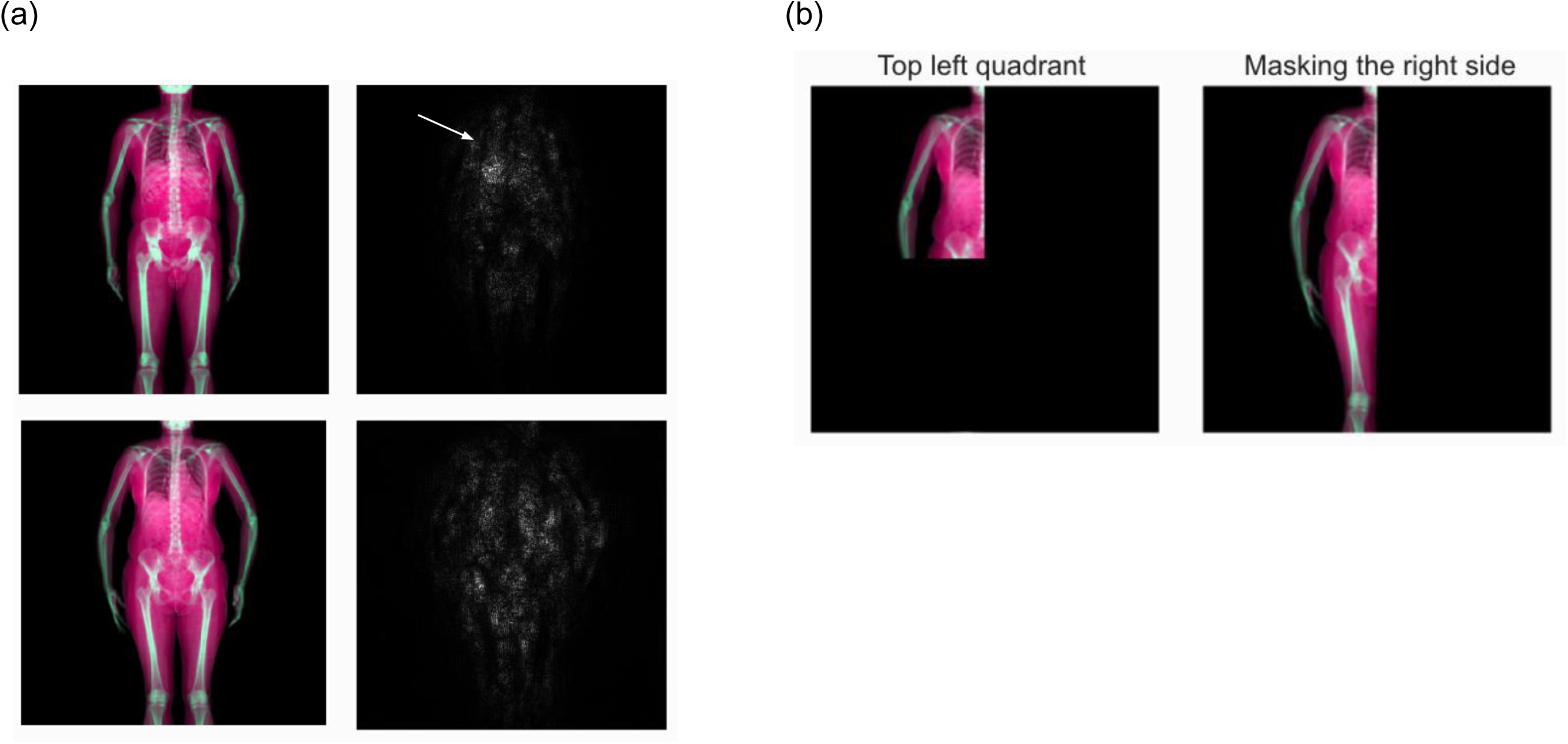
DXA saliency maps and masking experiments. (a) displays data from two individuals (rows). *Left panel*: DXA scans overlaid in the first two channels, with the third channel representing the mean of the first two. *Right panel*: shows their saliency maps, where pixel brightness is proportional to the calculated gradient. Gradients around the liver area are indicated by an arrow in the top left quadrant of the body. The top panel showcases data from a lean individual, while the bottom panel shows data from an individual with greater peripheral adiposity. (b) demonstrates example masking experiments within the DXA modality. *Left*: example showing subsetting the images to the top left quadrant. *Right*: example illustrating masking the right side of the image.

**Supplementary Fig. 4:**
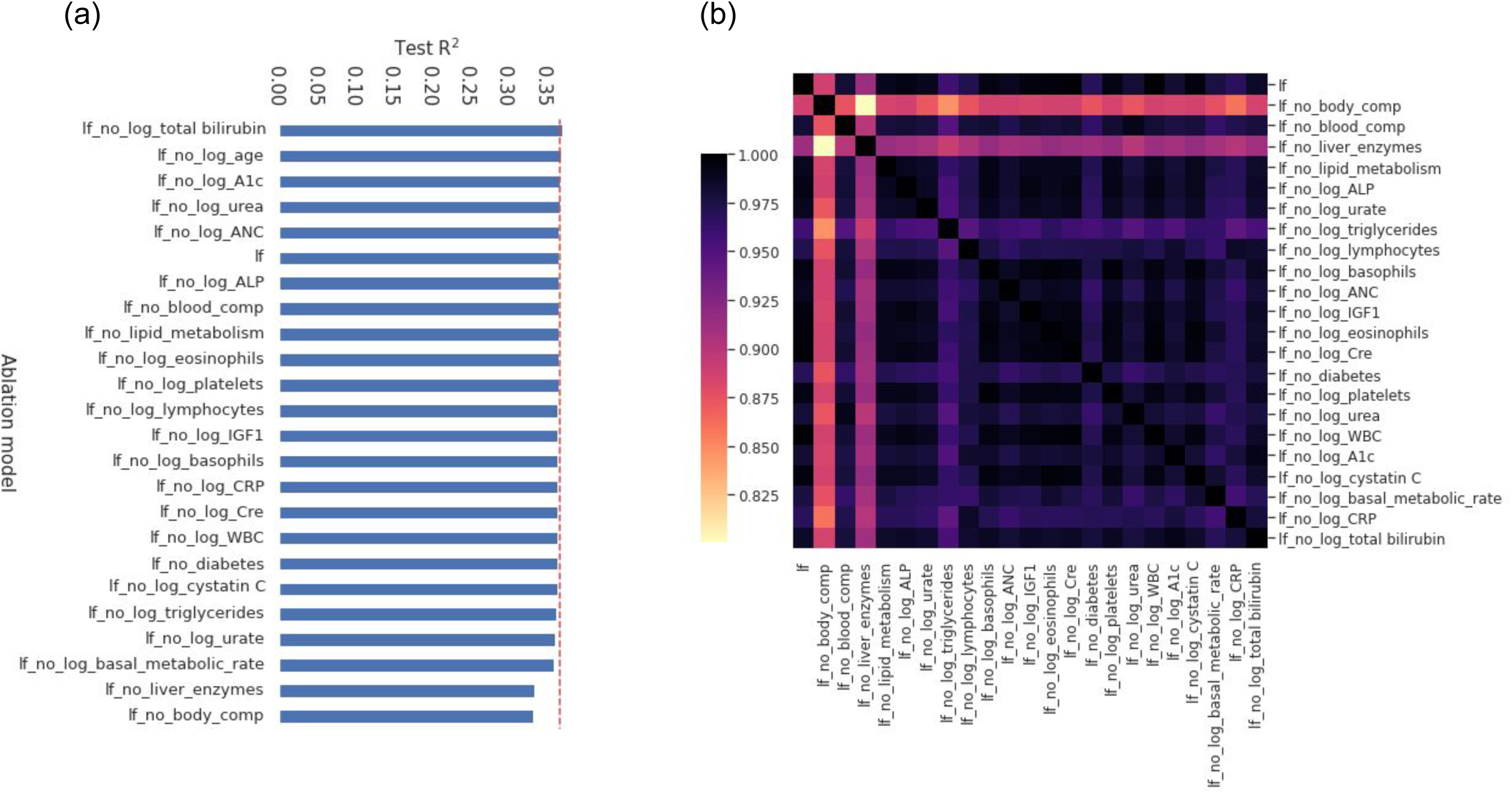
Results of the ablation analysis in the biomarker modality. (a) Performance (Pearson’s R^2^; *x* axis) of various ablation models (*y* axis) along with the main (unablated) model (“lf”) in the test set is shown. The dotted red line represents the R^2^ value from the main model. (b) Heatmap demonstrating the pairwise phenotypic correlation between liver fat predicted across the ablated models

**Supplementary Fig. 5:**
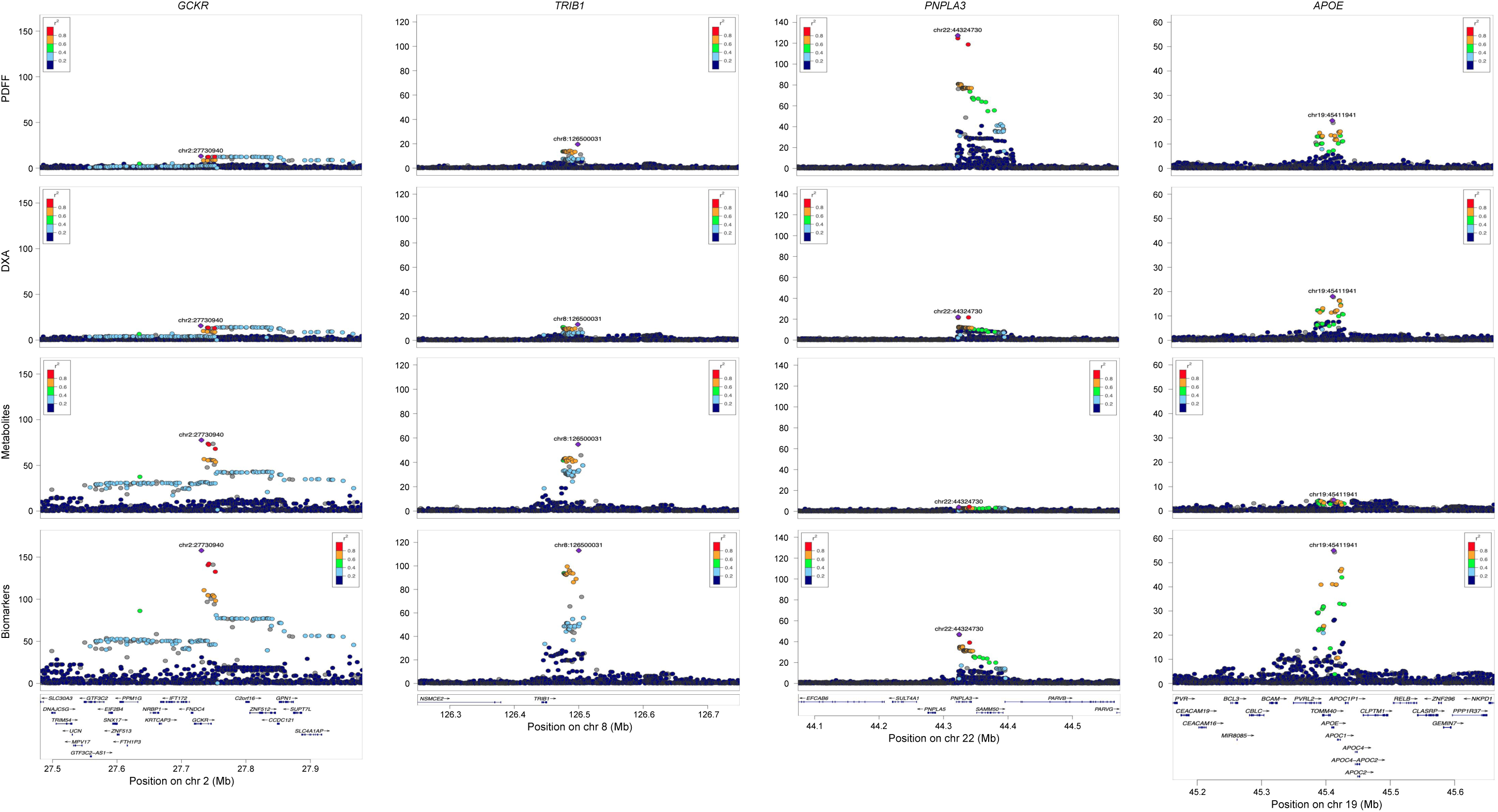
Regional association plots across modalities for examples of known loci.

**Supplementary Fig. 6:**
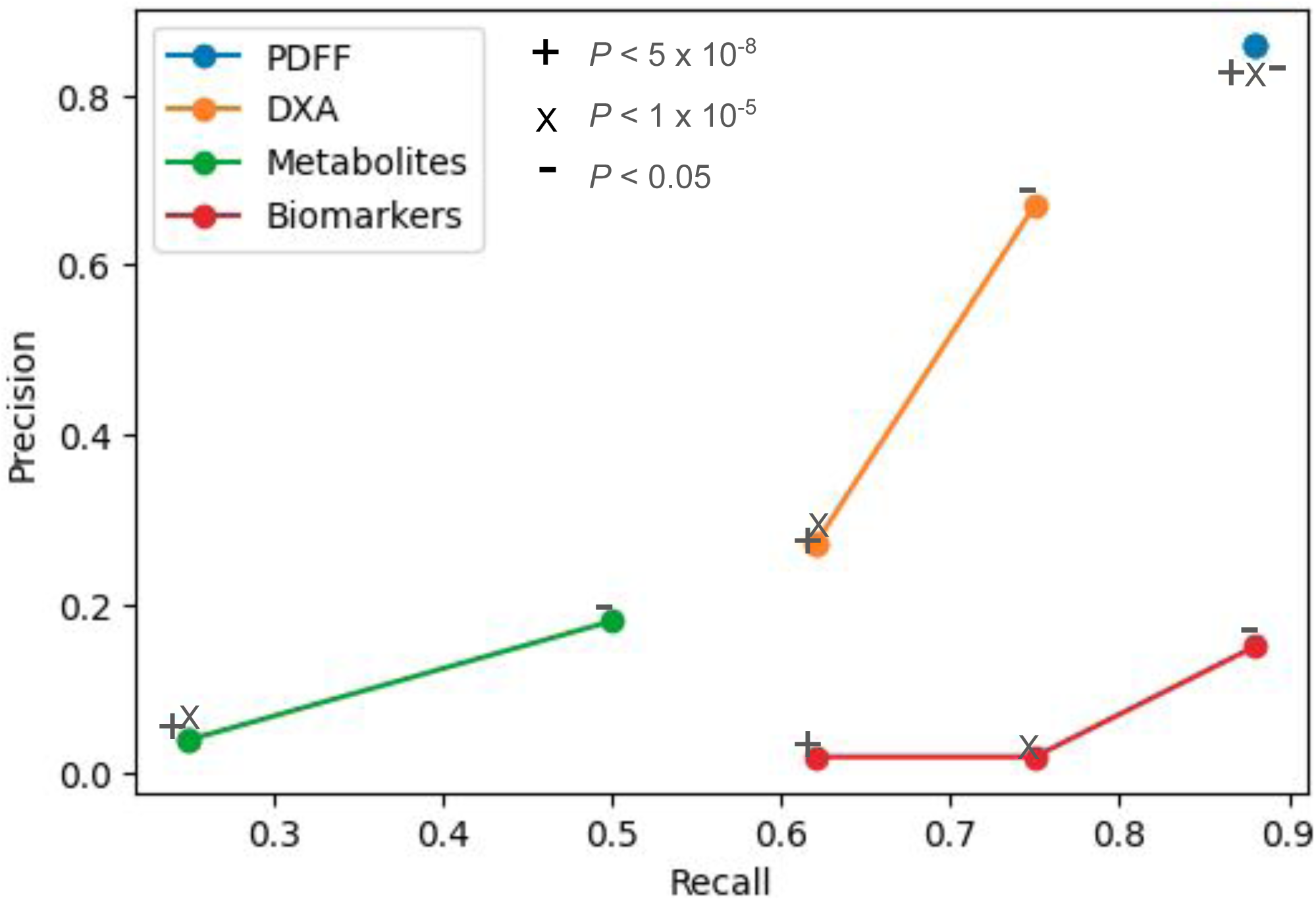
Precision-recall curves. Precision is plotted on the *y* axis, and recall on the *x* axis. Precision and recall were evaluated at various *P* value thresholds, including genome-wide significance (*P* < 5 x 10^-8^) indicated by ‘+’, suggestive significance (*P* < 1 x 10^-5^) indicated by ‘X’, and nominal significance (*P* < 0.05) indicated by ‘-’

**Supplementary Fig. 7:**
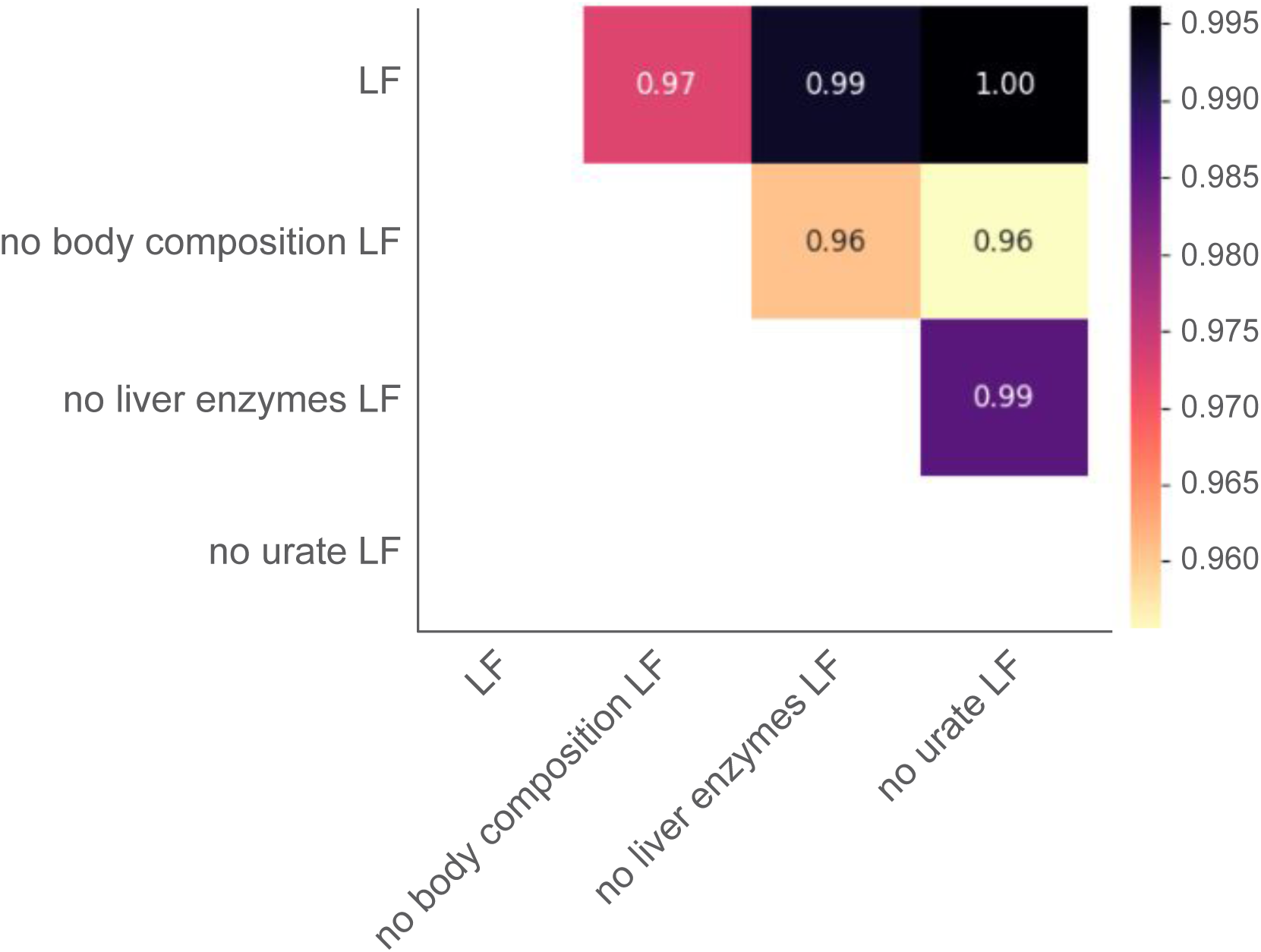
Genetic correlations across ablated phenotypes. Heatmap illustrating the pairwise genetic correlation estimates (*r*_g_) among the three ‘anomalous’ ablation sets and the main (unablated) predictions within the biomarker modality

**Supplementary Fig. 8:**
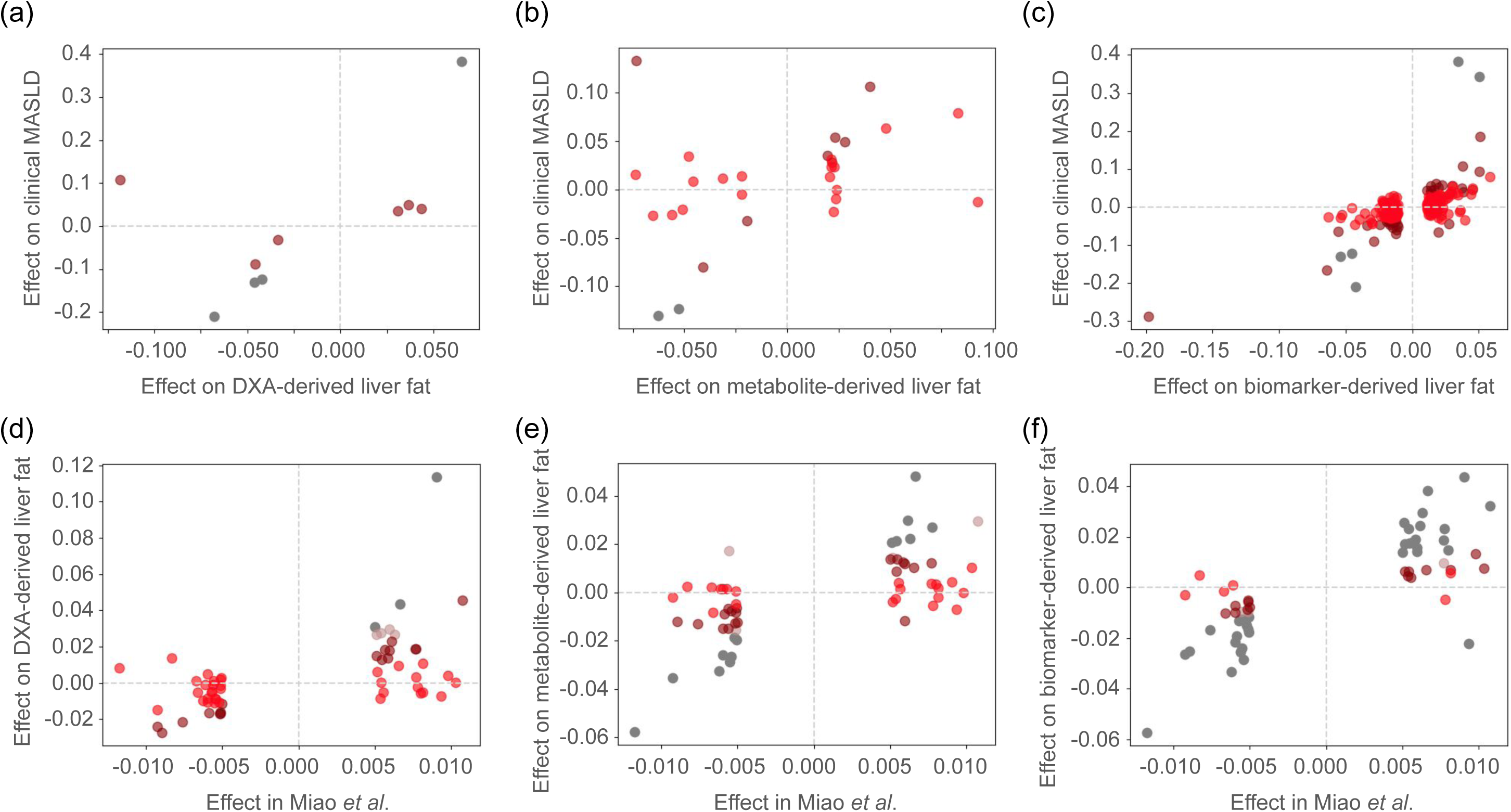
Directionally concordant effect of liver fat loci on clinical MASLD (**a - c**) and ML imputed MASLD status (Miao *et al*.; **d** - **f**). (**a**) Estimated effects between DXA-derived liver fat (*x* axis) and clinically diagnosed MASLD GWAS (*y* axis) at identified using DXA (at genome-wide significance). Loci for which data was present in the clinical MASLD GWAS are shown (n = 10). (**b**) Effect estimates between metabolite-derived liver fat (*x* axis) and clinical MASLD (*y* axis) at loci identified using metabolites. (**c**) Effect estimates between biomarker-derived liver fat (*x* axis) and clinical MASLD (*y* axis) at loci identified using biomarkers. (d - f) Estimated effects between Miao *et al* (*x* axis) and DXA-derived liver fat (*y* axis; **d**) metabolite-derived liver fat (*y* axis; **e**) and biomarker-derived liver fat (*y* axis; **f**) at genome-wide significant loci in Miao *et al*.. Loci are color coded based on their *P* value of association in clinical MASLD GWAS (**a** - **c**) and ML imputed MASLD GWAS (**d** - **e**): gray indicate *P* < 5 x 10^-8^; rosybrown indicate *P* < 1 x 10^-5^ and > 5 x 10^-8^; maroon indicate *P* < 0.05 and > 1 x 10^-5^; red indicate *P* > 0.05

